# Circulating microRNAs from early childhood and adolescence are associated with pre-diabetes at 18 years of age in women from the PMNS cohort

**DOI:** 10.1101/2021.11.24.21266835

**Authors:** Mugdha V. Joglekar, Pooja S. Kunte, Wilson K.M. Wong, D. S. Bhat, Sarang N. Satoor, Rohan R. Patil, Mahesh S. Karandikar, Caroline H.D. Fall, Chittaranjan S. Yajnik, Anandwardhan A. Hardikar

**Affiliations:** Diabetes and Islet Biology group, School of Medicine, Western Sydney University, Campbelltown, NSW 2560, Australia; Diabetes Unit, KEM Hospital and Research Center, Rasta Peth, Pune 411001, India; National Centre for Cell Science, DNA Sequencing facility, NCMR Campus, Level 3, Sai Trinity Building, Central Tower, Pashan-Sus Road, Pashan, Pune 411026, India; DY Patil Medical College, DY Patil University, Pimpri, Pune, 411048; MRC Lifecourse Epidemiology Unit, Southampton University and General Hospital, Southampton, UK

**Keywords:** MicroRNAs, pre-diabetes, Pune Maternal Nutrition Study, machine learning

## Abstract

A high (20%) prevalence of glucose intolerance at 18-years was seen in women from the Pune Maternal Nutrition Study (PMNS) birth cohort. Here, we provide preliminary longitudinal analyses of circulating microRNAs in normal glucose tolerant (NGT@18y, N=10) and glucose intolerant (N=8) women (ADA criteria) at 6-, 12- and 17-years of their age using discovery analysis (OpenArray™ platform). Machine-learning workflows involving Lasso with bootstrapping/leave-one-out cross-validation (LOOCV) identified microRNAs associated with glucose intolerance at 18-years of age. Several microRNAs, including miR-212-3p, miR-30e-3p and miR-638, stratified glucose-intolerant women from NGT at childhood. Our results suggest that circulating microRNAs in childhood could predict pre-diabetes at 18-years of age. Validation of these findings in males and remaining participants from the PMNS birth cohort will provide a unique opportunity to study novel epigenetic mechanisms in the life-course progression of glucose intolerance and enhance current clinical risk prediction of pre-diabetes and progression to type 2 diabetes.

## INTRODUCTION

Pre-diabetes includes hyperglycaemia in the fasting state (impaired fasting glucose/IFG) and/or hyperglycaemia after feeding/postprandial state (impaired glucose tolerance/IGT). Current diagnosis of pre-diabetes is based on family history of diabetes, overweight/obesity, and glucose testing, either as an oral glucose tolerance test (OGTT)^1^, or HbA1c measurement (ADA clinical practice). The prevalence of pre-diabetes is increasing globally, and undetected pre-diabetes is high-risk for the development of future T2D and cardiovascular diseases. Screening tools involving biochemical and/or molecular biomarkers may improve pre-diabetes prediction and help in early risk-stratification of individuals via lifestyle intervention^2, 3^.

In 1993, a birth cohort was set up in rural Pune, India with the aim of understanding *in utero* determinants of fetal growth and life-course evolution of the phenotype for diabetes and related disorders. This unique cohort, referred to as the Pune Maternal Nutrition Study (PMNS), has plasma samples and clinical information from around 700 families over three generations (F_0_, F_1_, F_2_) with anthropometric and biochemical measurements in parents and offspring^4^. We observed that 20% of F_1_ daughters born to F_0_ mothers developed pre-diabetes at 18 years of age^5^. The serial database and biorepository of samples from PMNS provided a unique opportunity to test the predictive value of early life (6-,12-, 17-years) circulating microRNAs in the development of pre-diabetes at a young age.

MicroRNAs are 18-22 nucleotide, regulatory non-coding RNAs that can bind to protein-coding gene transcripts (mostly at the 3’UTR) and either degrade or render these mRNA transcripts translationally inactive^6^. The regulatory and mechanistic role of microRNAs in physiological and pathological processes associated with diabetes remains to be discovered. Circulating microRNAs offer the potential to serve as minimally invasive biomarkers of diabetes-associated pathophysiological features^7^, enabling the prediction of disease at an earlier stage. Considering the importance of early detection of pre-diabetes as well as the need for additional biomarkers to improve the accuracy of pre-diabetes prediction, we aimed to study microRNAs in the PMNS cohort. We profiled all known/validated (N=754) microRNAs in F_1_ daughters at 6-, 12- and 17-years of age to identify differentially expressed microRNAs associated with the clinical diagnosis of pre-diabetes at 18 years of age. Our data identify microRNAs associated with and potentially predictive of future pre-diabetes and provide the first report of longitudinal changes in circulating microRNAs, as biomarkers of metabolic disease in later life.

## METHODS

### Cohort description

The PMNS birth cohort was set up in six villages near Pune, India^4^. A total of 797 pregnant women (F_0_) were enrolled along with their spouses and studied three times during the pregnancy. F_1_ children were assessed at birth, 6-, 12-, 17- and 18-years of age for anthropometric, biochemical and blood glucose measurements. Details including dietary intake, physical activity and socio-economic status were also noted. The study was approved by the King Edward Memorial (KEM) Hospital Research Centre, Pune and followed the Indian Council for Medical Research (ICMR), Government of India guidelines for the ethical conduct of the study. All study participants signed informed consent at 18 years of follow up; at 6, 12 and 17-years, parents signed informed consent, and at 12 and 17-years, children also signed an assent.

### Glucose measurements

An oral glucose tolerance test (OGTT, 1.75g/kg of anhydrous glucose) was performed at 6-years and a 75g OGTT at 18 years of age. Fasting, 30-mins, 120-mins blood samples were collected during the OGTT. At 12-years, only a fasting blood sample and at 17-years a random blood sample was collected. Plasma was separated by centrifugation at 1000xg for 10 min and stored at -80°C. Blood glucose was measured by the glucose oxidase/peroxidase method.

### Diagnosis of Pre-diabetes

At 18 years of age glucose tolerance was classified by ADA criteria (www.diabetes.org), as normal glucose tolerant (NGT) if fasting plasma glucose (FPG)<100 mg/dL and 2-hour plasma glucose (PG) <140 mg/dL); or as impaired fasting glucose (IFG) if FPG 100-125 mg/dL and 2 hour PG < 140 mg/dL, as impaired glucose tolerant (IGT) if FPG < 100 mg/dL and 2 hour PG 140-199 mg/dL. Pre-diabetes includes IFG and IGT. For this study, we selected female participants with pre-diabetes (N=8) and NGT (N=10).

### RNA isolation

Stored plasma samples (100-200µl) from all participants and at all three time-points (6-, 12- and 17-years of age) were used for RNA isolation as detailed earlier^8, 9^. The automation involved isolation of total RNA using the QIAcube-HT platform (Qiagen, Hilden, Germany) as described^9^. Briefly, 500µl of TRIzol reagent (ThermoFisher Scientific, USA) and 10ng of glycogen (Sigma-Aldrich, Hamburg, Germany) were added to each sample. The aqueous phase was then separated following centrifugation and then used for RNA purification using the RNeasy-HT Kit (Qiagen, Hilden, Germany) on an automated platform. Manual steps involved isopropanol addition for RNA precipitation followed by washing with freshly prepared 75% ethanol. RNA was dissolved in nuclease-free water and stored at -80°C until further analysis. RNA concentration/purity was assessed using a NanoDrop^™^ spectrophotometer (ThermoFisher Scientific, USA) before proceeding for microRNA qPCR.

### MicroRNA measurement using OpenArray PCR

RNA was converted to cDNA using Megaplex Human RT Primers (pool A and pool B) and TaqMan® microRNA RT Kit (both from ThermoFisher Scientific, USA). A 10ng input RNA was used with the manufacturer’s “low sample input (LSI)” protocol for reverse transcription, pre-amplification and real-time qPCR. Pre-amplification was performed on the entire cDNA product using Megaplex^™^ PreAmp Primers and TaqMan PreAmp Master Mix (both from ThermoFisher Scientific, USA) for 16 cycles. The pre-amplified product was diluted 1:20 in 0.1×TE buffer (pH 8.0) and then mixed with TaqMan^™^ OpenArray^®^ PCR master mix before loading onto TaqMan OpenArray^®^ Human microRNA inventoried panels using AccuFill™ robotic system (Life Technologies, Foster City, CA, USA). The discovery set of 754 microRNAs was measured in samples from all women (N=10 NGT and N=8 pre-diabetes X 3 time points) using TaqMan-based RT-qPCR with OpenArray^®^ platform on QuantStudio 12K Flex System (Life Technologies, Foster City, CA, USA) as described earlier^10^. Data were imported to ThermoConnect software and normalised using a built-in global normalisation method after removing any undetectable microRNAs (presenting amplification score <1.24 and cycle value (Cq) confidence interval <0.6). Normalized Ct-values were exported to a CSV format for analysis. MicroRNAs considered undetected/not expressed were assigned a Ct-value of 39, which corresponds to the limit of detection for the TaqMan™ qPCR system in our hands^11,12^. Results are calculated using the ΔCt method (difference in normalised Ct-values between pre-diabetes progressors and non-progressors (i.e. NGT)).

### Data analysis and Statistics

To reliably measure a two-fold change in the levels of microRNAs relative to NGTs (the equivalent of one Ct-value difference by qPCR), and assuming a standard deviation (SD) of 30% of the mean (effect size=1.667^13^) with α=0.05, power=87%, we would need 8 individuals per group. With samples from 8 pre-diabetes and 10 NGT participants at all three time points, the observed SDs were much smaller (<13% of mean; all time points) than the expected 30%, thereby offering the desired statistical power for these analyses. Data were checked for normal distribution before selecting a parametric or non-parametric test. Demographic characteristics of the study participants are represented as z-scores and significance was calculated by t-test. Z-score standardization for each variable in pre-diabetes was calculated relative to NGT as per the formula: (mean(pre-diabetes[i])-mean(NGT[i]))/SD(NGT[i]), where i=clinical characteristic. MicroRNAs that were undetected (Ct-value ≥39) across all samples in both groups were removed from the analysis. Penalized logistic regression using least absolute shrinkage and selection operator (LASSO) and bootstrapping was employed to identify microRNAs associated with pre-diabetes at 18 years. Two different R-scripts were used by separate investigators on deidentified datasets. Through these approaches, microRNAs that had a non-zero β-coefficient and a non-zero frequency were selected for Receiver Operating Characteristic (ROC) curve analysis. Fasting blood glucose was also considered as another variable in the ROC curve analysis, which was performed using leave-one-out cross-validation (LOOCV) approach with an in-house R pipeline and trainControl (Caret, version 6.0). Prediction performance was assessed by calculating sensitivity, specificity, positive predictive value, negative predictive value, and accuracy from confusion matrix. Area under the curve (AUC) was calculated using pROC (v1.16.2) function as described earlier^9^ with R software ver. 3.6.2 (R Foundation for Statistical Computing, Vienna, Austria). A categorical bubble plot was generated using the R packages ggplot2 (3.3.2) and ggpubr (0.4.0) as described earlier^14^. Forcats package (0.5.1) was included to arrange variables (presented on y-axis) in order.

### Pathway analysis

Kyoto Encyclopaedia of Genes and Genomes (KEGG) pathway analysis was carried out using a web tool (miRSystem, ver. 20160513; http://mirsystem.cgm.ntu.edu.tw/) for each set of important microRNAs identified at 6-, 12- and 17-years, associated with pre-diabetes at 18-years of age, using methodologies described earlier^9, 14^. Significant raw p-values (≤0.05) of the KEGG pathways are considered.

## RESULTS

### Clinical characteristics of PMNS participants

Of the 213 (80%) NGT women and 54 (20%) presenting with pre-diabetes at 18 years of age, 10 NGT and age-matched eight women with pre-diabetes were selected for this study. **Figure 1A** presents their clinical and biochemical characteristics at all ages. There was no significant difference in the BMI of the two groups at 18-, 17-, 12- and 6-years of age. By definition, at 18-years, women with pre-diabetes had higher glucose concentrations during the OGTT, and lower insulin secretion with lower insulin sensitivity. In this small subset of PMNS participants, there was no significant difference in fasting plasma glucose (FPG) compared to NGT (P>0.05) at 6- and 12-years. Other measurements including fasting insulin, HOMA indices as well as skinfold thickness were also similar between the participants at earlier (−6, -12, -17 years) ages. (**Figure 1A**).

**Figure 1:**
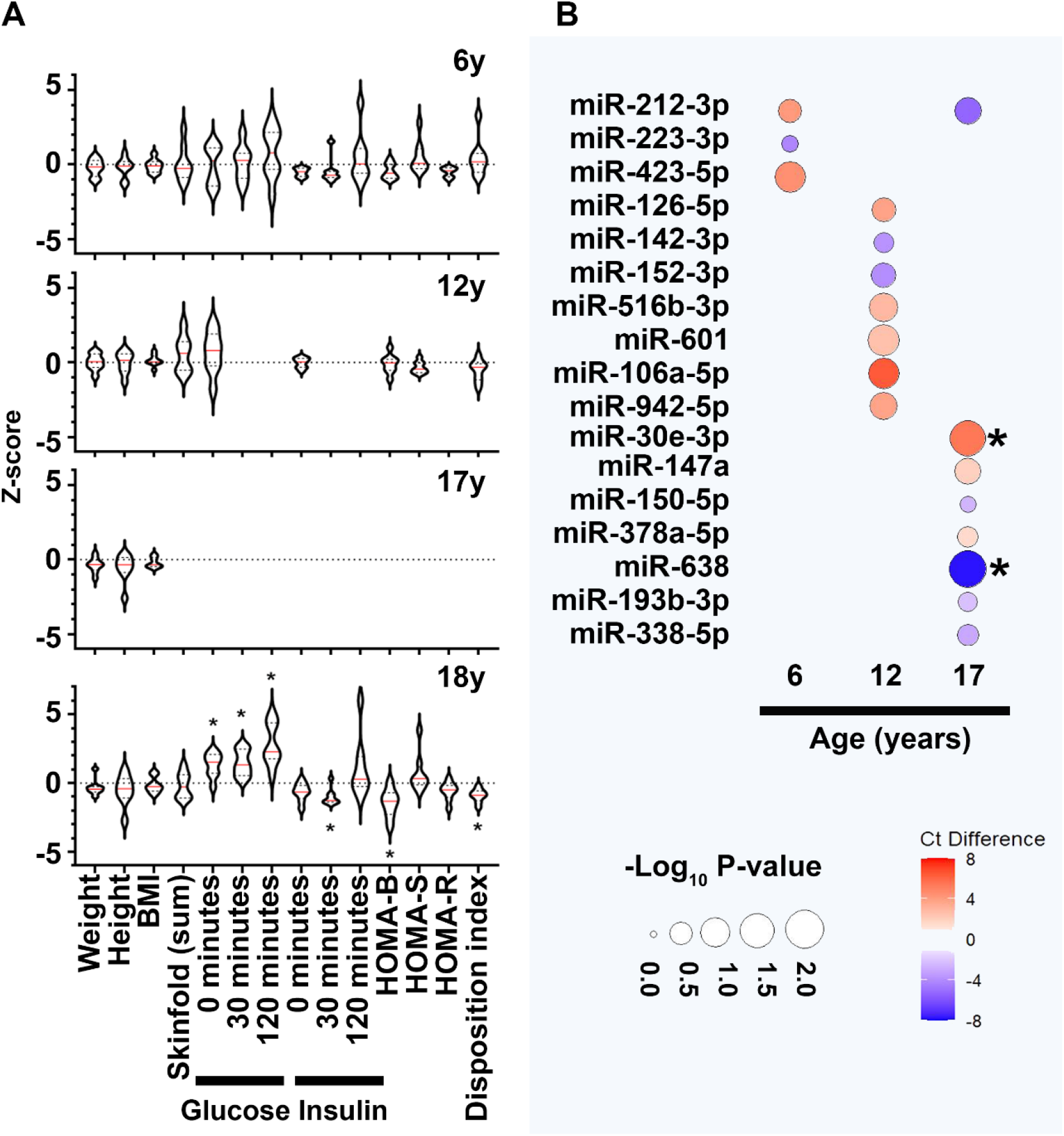
Clinical characteristics and dysregulated microRNAs in PMNS study samples A) Data presented as z-score for the respective clinical measure presented on the X-axis. The Z-score is calculated for the participants with pre-diabetes at 18 years (N=8) relative to the mean of NGT (N=10) group at each time point. Violin plots display the median (red line) within each polygon violin and each polygon represents the density of the data and extends to min/max values. Significance is calculated using a t-test and *=p<0.05. B) Categorical bubble plot displaying the average Ct-value difference of the important microRNAs (identified via penalized logistic regression with bootstrap for NGT vs pre-diabetes analysis); listed on Y-axis) across age (presented on X-axis). The average Ct-value difference was calculated as (Ct-value of NGT – Ct-value of pre-diabetes) and presented in colours ranging from red to blue. Red indicates lower average microRNA abundance; while blue indicates higher average microRNA abundance in NGT compared to pre-diabetes group. The p-value is presented by the size of the bubble, with larger circles presenting higher significance. The microRNAs demonstrating significant differences (p<0.05) are marked with (*) next to their bubble. The p-values were calculated with Mann-Whitney test.

### Profiling of microRNAs at earlier ages identified important biomarkers of pre-diabetes

Discovery profiling of microRNAs in women with pre-diabetes or NGT identified up to 161 microRNAs that were detectable in at least one of the study samples at any given age. This is in accordance with our previous studies^9, 12^ where we have observed a smaller number of microRNAs in circulation compared to those within cellular compartments. Bootstrapping approach offered minimizing sampling bias and ranked microRNAs on their importance (bootstrap frequency) for each of the time points assessed. A total of 17 different microRNAs with non-zero β-coefficient and a non-zero frequency in penalized logistic regression using LASSO and bootstrapping were identified in 6-, 12-, and 17-year samples. Three microRNAs at 6-years, seven at 12-years and eight at 17-years associated with future pre-diabetes (**Figure 1B**); miR-212-3p was common amongst the microRNAs identified at 6- and 17-years for their association with pre-diabetes at 18-years (**Figure 1B**).

### MicroRNAs offered better risk-stratification for future pre-diabetes than blood glucose

Fasting glucose is an important clinical predictor of future pre-diabetes^15^. We, therefore performed ROC curve analyses to assess if microRNAs offered any predictive advantage over fasting glucose concentrations at 6- and 12-years, in stratifying NGT and pre-diabetes at 18 years of age. At 6-years of age, three of the microRNAs (miR-212-3p, -223-3p, -423-5p) together offered a higher prediction accuracy (accuracy: 88%, AUC: 0.82) than individual microRNAs (accuracy: 66% to 72%, AUC: 0.37 to 0.51) or fasting glucose alone (accuracy: 38%, AUC: 0.0) (**Table 1**). Fasting glucose alone at 6 years, could not identify the true progressors. Similarly, at 12-years, seven different microRNAs together performed better (accuracy: 77%, AUC=0.89) than individual microRNAs (accuracy: 55% to 72%, AUC: 0.34 to 0.63) and fasting glucose alone (accuracy: 55%, AUC=0.5) (**Table 1**). At each age-group, combined models using all selected/important microRNAs offered better risk-stratification (higher ROC AUC) than glucose alone (**Table 1**). Pathway analysis using the important microRNAs indicated TGF-β signalling as a common targeted pathway at all age groups (Fig S1).

**Table 1:** ROC curve analysis ROC curve analysis was performed using Leave-One-Out Cross Validation (LOOCV) approach. Variables include the important microRNAs identified (in NGT vs pre-diabetes analysis) at each age (6- and 12-years) along with fasting blood glucose. Specificity, sensitivity, AUC, positive predictive value, negative predictive value, accuracy with 95% confidence interval (CI) for each microRNA alone, fasting glucose alone and their combinations is shown.

|  | Specificity | Sensitivity | AUC | Pos Pred Value | Neg Pred Value | Accuracy | 95% CI |
| --- | --- | --- | --- | --- | --- | --- | --- |
| **6Y** |  |  |  |  |  |  |  |
| miR-212 | 0.80 | 0.50 | 0.40 | 0.66 | 0.66 | 0.66 | (0.41, 0.87) |
| miR-223 | 0.80 | 0.50 | 0.51 | 0.66 | 0.66 | 0.66 | (0.41, 0.87) |
| miR-423-5p | 1.00 | 0.38 | 0.38 | 1.00 | 0.66 | 0.72 | (0.47, 0.90) |
| fasting glucose | 0.70 | 0.00 | 0.00 | 0.00 | 0.46 | 0.38 | (0.17, 0.64) |
| all miRs | 0.90 | 0.87 | 0.83 | 0.88 | 0.90 | 0.88 | (0.65, 0.99) |
| miRs+glucose | 0.90 | 0.75 | 0.81 | 0.86 | 0.82 | 0.83 | (0.59, 0.96) |
| **12Y** |  |  |  |  |  |  |  |
| miR-126# | 0.60 | 0.50 | 0.45 | 0.50 | 0.60 | 0.55 | (0.31, 0.79) |
| miR-142-3p | 0.80 | 0.63 | 0.50 | 0.71 | 0.73 | 0.72 | (0.47, 0.90) |
| miR-152 | 0.50 | 0.88 | 0.44 | 0.58 | 0.83 | 0.66 | (0.41, 0.87) |
| miR-516-3p | 0.80 | 0.63 | 0.50 | 0.71 | 0.73 | 0.72 | (0.47, 0.90) |
| miR-601 | 0.70 | 0.50 | 0.63 | 0.57 | 0.64 | 0.61 | (0.36, 0.83) |
| miR-106a | 1.00 | 0.38 | 0.38 | 1.00 | 0.66 | 0.72 | (0.47, 0.90) |
| miR-942 | 0.90 | 0.38 | 0.34 | 0.75 | 0.64 | 0.66 | (0.41, 0.87) |
| fasting glucose | 0.70 | 0.38 | 0.50 | 0.50 | 0.58 | 0.55 | (0.31, 0.79) |
| all miRs | 0.70 | 0.88 | 0.89 | 0.70 | 0.88 | 0.77 | (0.52, 0.94) |
| miRs+glucose | 0.60 | 0.63 | 0.63 | 0.56 | 0.67 | 0.61 | (0.36, 0.83) |

## DISCUSSION

This novel and preliminary report presents a longitudinal analysis of microRNA expression in women with or without pre-diabetes at 18-years. In this small subset of randomly selected PMNS women, we identified important microRNAs at 6-, 12- and 17-years of age in women with pre-diabetes (vs NGT), although clinical features, including plasma glucose concentrations were similar. The identification of microRNAs that are associated with and potentially predictive of pre-diabetes in later life could help in efficient risk-stratification and better management of individuals at risk of diabetes.

Several candidates such as proteins, metabolites and microbiota have been tested for their prognostic/diagnostic capacity to predict pre-diabetes^1,16,17^. Systematic review and meta-analyses indicate that circulating concentrations of metabolites, including hexoses, amino acids, phospholipids, and triglycerides are associated with pre-diabetes^18^.

MicroRNAs are an interesting class of biomarkers as they also present the capacity to be regulators of underlying pathophysiological mechanisms leading to pre-diabetes. In a recent, large cross-sectional study, miR-30e-3p and miR-126-3p were found to be highly abundant and consistently associated with pre-diabetes^19^. Another study reported differentially expressed microRNAs (including miR-223-3p) at baseline in pre-diabetes individuals who later progressed to T2D^20^. In samples from the ORIGINS trial, multiple microRNAs including miR-126, miR-150, miR-223 were found to be significantly dysregulated in pre-diabetes and T2D^21^. MicroRNA-192 and -193b are reported to be significantly higher in pre-diabetes stage and return to lower levels following exercise^22^. The majority of microRNAs reported to be dysregulated in pre-diabetes through these studies^19-22^ are also seen to be differentially expressed in our analysis (including miR-30e, miR-223, miR-126, miR-150 and miR-193b) (**Figure 1B**).

TGFβ pathway ligands play an important role in endocrine pancreas function^23^, and the expression/abundance of TGFβ has been shown to increase following exposure to a high-fat diet^24^ in the fruit fly, and also increased in circulation in clinically observed pre-diabetes^25^. The increase in microRNAs targeting TGFβ pathways, observed at 6-, 12- and 17 years of age in this study, indicates a protective response through inhibitory microRNAs that target the TGFβ pathway. To the best of our knowledge, we are the first to report potential predictive microRNA candidates of pre-diabetes from early childhood, adolescence to adulthood in south Asian women. We observed that microRNA profiles tend to be different at early ages; however, the anthropometric and other clinical/biochemical characteristics at those time points were similar in the study participants. Samples are obtained from a well-characterized multigenerational birth cohort. We used real-time PCR based microRNA profiling for all known and validated microRNAs instead of selecting candidate microRNAs from previous publications. Such a discovery approach was important for unprejudiced identification of key microRNAs in our cohort as microRNAs may differ with ethnicity, age and sex^26^. We used unbiased machine learning analytical methods to remove sampling bias. Fasting glucose alone had low AUCs (0 at 6-years and 0.5 at 12-years) in ROC curve analysis (Table 1). Even though not significant in these smaller sample sets, FPG at 6- and 12-years is known to be a strong predictor of pre-diabetes at 18-years in the PMNS^5^. Overall, we think that the microRNAs identified in this preliminary report, along with clinical measurements will further enhance the prediction for onset of pre-diabetes in this cohort. The small sample size is a limitation; although, this was adequately powered to detect the desired effect and addressed by selecting appropriate (LOOCV) techniques for cross-validation. Validation in all remaining individuals (males; 221 NGT and 131 pre-diabetes, and females; 203 NGT and 46 pre-diabetes) is planned. This preliminary study also opens future research opportunities to test the potential of microRNAs in determining intergenerational (as well as trans-generational) traits and predisposition to pre-diabetes/T2D in later life.

## Data Availability

All data produced in the present study are available upon reasonable request to the authors

## Acknowledgements

We acknowledge participants and their families for supporting this study. Staff at Diabetes Unit for recruitment, patient follow-up and sample collection are greatly appreciated. Infrastructure support from KEM Hospital, D. Y. Patil University, University of Sydney, and Rebecca Cooper Medical Foundation is acknowledged.

## Financial Support: (include grant numbers)

Fellowship support from Australian Diabetes Society (Skip Martin Award, 2016) and AISRF EMCR fellowship 2018-19 (Australian Academy of Science) to MVJ is acknowledged. AAH is supported by grants from Juvenile Diabetes Research Foundation (JDRF) Australia T1D Clinical Research Network (JDRF/4-CDA2016-228-MB) and Visiting Professorships (2016-18 and 2019-22) from the Danish Diabetes Academy, funded by the Novo Nordisk Foundation, grant number NNF17SA0031406. WKMW is supported through the Leona M. and Harry B. Helmsley Charitable Trust (Grant 2018PG-T1D009) in collaboration with the JDRF Australian Type 1 Diabetes Clinical Research Network funding (Grant 3-SRA-2019-694-M-B to AAH). PMNS was funded by the Wellcome Trust, UK (038128/Z/93, 059609/Z/99, 079877/Z/06/Z, 098575/B/12/Z and 083460/Z/07/Z), MRC, UK (MR/J000094/1) and Department of Biotechnology, GoI (BT/PR-6870/PID/20/268/2005). Between these grants, the study was funded intramurally (KEMHRC). CSY and AAH were both visiting professors to the Danish Diabetes Academy when these measurements were made.

## Ethical Standards

The study was approved by village leaders and the KEM Hospital Research Centre Ethics Committee. Parents gave written consent; children under 18 years of age gave written assent, and written consent after reaching 18 years.

## Author contributions

Experimental work: MVJ, SNS, RRP, MSK, AAH; Data analysis: MVJ, PSK, WKMW; Sample collection: DB, CSY; Writing: MVJ, PSK, WKMW, CSY, AAH; Conceptualisation and funding: CSY, CF, AAH.

**Supplementary Figure 1:**
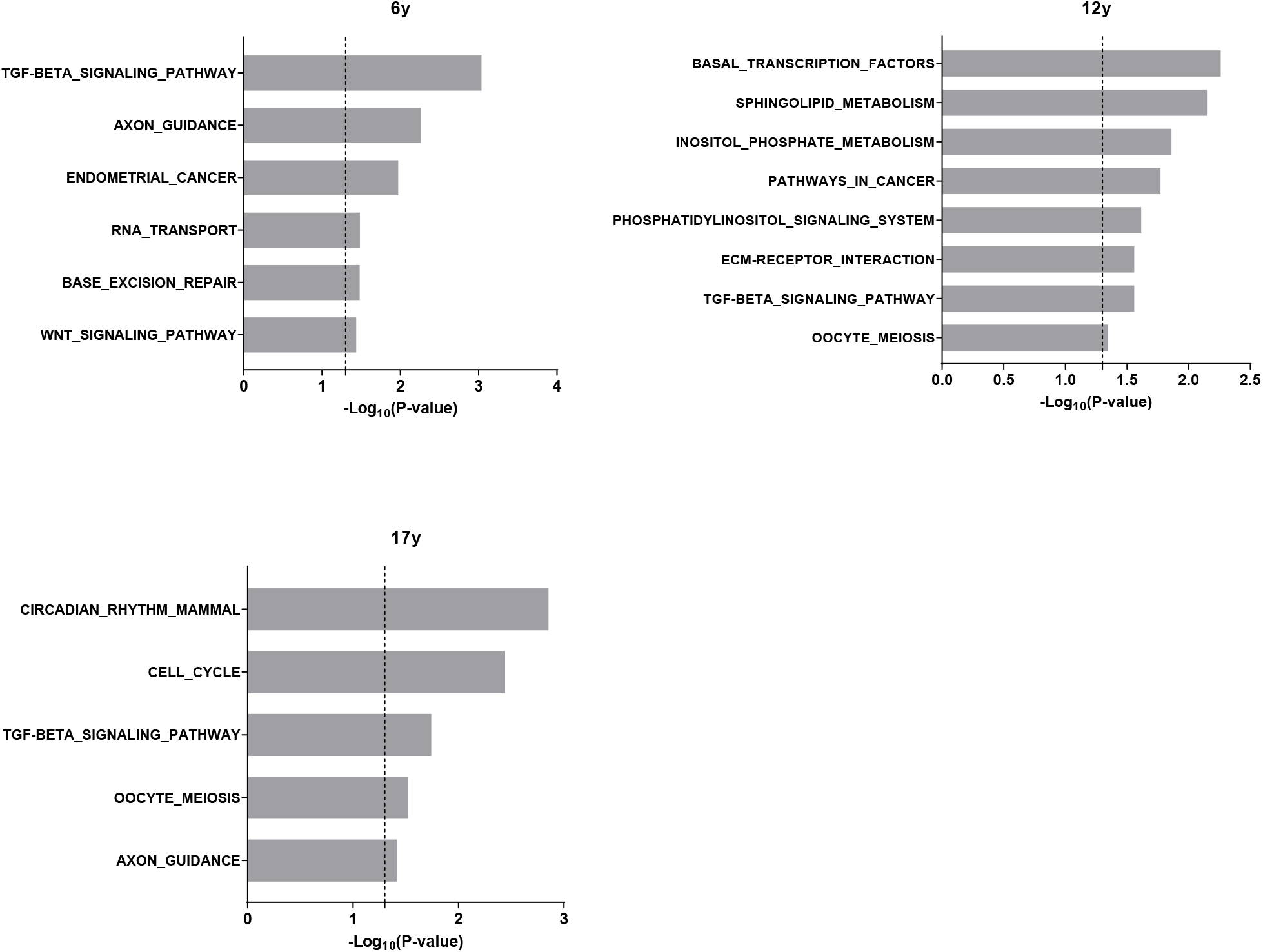
Pathway analysis Significant KEGG pathways associated with the important microRNAs identified (in NGT vs pre-diabetes analysis) at each age (6-, 12- and 17-years).

## References

1. Bergman M, Abdul-Ghani M, DeFronzo RA, et al. Review of methods for detecting glycemic disorders. Diabetes Res Clin Pract. 2020;165, 108233.

2. Bell K, Shaw JE, Maple-Brown L, et al. A position statement on screening and management of pre-diabetes in adults in primary care in Australia. Diabetes Res Clin Pract. 2020;164, 108188.

3. Campbell MD, Sathish T, Zimmet PZ, et al. Benefit of lifestyle-based T2DM prevention is influenced by pre-diabetes phenotype. Nat Rev Endocrinol. 2020;16(7), 395–400.

4. Rao S, Yajnik CS, Kanade A, et al. Intake of micronutrient-rich foods in rural Indian mothers is associated with the size of their babies at birth: Pune Maternal Nutrition Study. J Nutr. 2001;131(4), 1217–1224.

5. Yajnik CS, Bandopadhyay S, Bhalerao A, et al. Poor <em>in-utero</em> growth, and reduced beta cell compensation and high fasting glucose from childhood, are harbingers of glucose intolerance in young Indians. 2021; doi: 10.1101/2020.11.19.20234542 %J medRxiv, 2020.2011.2019.20234542.

6. Wong WKM, Sorensen AE, Joglekar MV, Hardikar AA, Dalgaard LT. Non-Coding RNA in Pancreas and beta-Cell Development. Noncoding RNA. 2018;4(4).

7. Guay C, Regazzi R. Circulating microRNAs as novel biomarkers for diabetes mellitus. Nat Rev Endocrinol. 2013;9(9), 513–521.

8. Taylor CJ, Satoor SN, Ranjan AK, Pereira e Cotta MV, Joglekar MV. A protocol for measurement of noncoding RNA in human serum. Exp Diabetes Res. 2012;2012, 168368.

9. Joglekar MV, Wong WKM, Ema FK, et al. Postpartum circulating microRNA enhances prediction of future type 2 diabetes in women with previous gestational diabetes. Diabetologia. 2021; doi: 10.1007/s00125-021-05429-z.

10. Farr RJ, Januszewski AS, Joglekar MV, et al. A comparative analysis of high-throughput platforms for validation of a circulating microRNA signature in diabetic retinopathy. Sci Rep. 2015;5, 10375.

11. Hardikar AA, Farr RJ, Joglekar MV. Circulating microRNAs: understanding the limits for quantitative measurement by real-time PCR. J Am Heart Assoc. 2014;3(1), e000792.

12. Shihana F, Joglekar MV, Raubenheimer J, Hardikar AA, Buckley NA, Seth D. Circulating human microRNA biomarkers of oxalic acid-induced acute kidney injury. Arch Toxicol. 2020;94(5), 1725–1737.

13. Olejnik S, Algina J. Generalized eta and omega squared statistics: measures of effect size for some common research designs. Psychol Methods. 2003;8(4), 434–447.

14. Wong WKM, Joglekar MV, Saini V, et al. Machine learning workflows identify a microRNA signature of insulin transcription in human tissues. iScience. 2021;24(4), 102379.

15. Feizi A, Meamar R, Eslamian M, Amini M, Nasri M, Iraj B. Area under the curve during OGTT in first-degree relatives of diabetic patients as an efficient indicator of future risk of type 2 diabetes and pre-diabetes. Clin Endocrinol (Oxf). 2017;87(6), 696–705.

16. Vaishya S, Sarwade RD, Seshadri V. MicroRNA, Proteins, and Metabolites as Novel Biomarkers for Pre-diabetes, Diabetes, and Related Complications. Front Endocrinol (Lausanne). 2018;9, 180.

17. Zhou W, Sailani MR, Contrepois K, et al. Longitudinal multi-omics of host-microbe dynamics in pre-diabetes. Nature. 2019;569(7758), 663–671.

18. Guasch-Ferre M, Hruby A, Toledo E, et al. Metabolomics in Pre-diabetes and Diabetes: A Systematic Review and Meta-analysis. Diabetes Care. 2016;39(5), 833–846.

19. Weale CJ, Matshazi DM, Davids SFG, et al. MicroRNAs-1299, -126-3p and -30e-3p as Potential Diagnostic Biomarkers for Pre-diabetes. Diagnostics (Basel). 2021;11(6).

20. Parrizas M, Mundet X, Castano C, et al. miR-10b and miR-223-3p in serum microvesicles signal progression from pre-diabetes to type 2 diabetes. J Endocrinol Invest. 2020;43(4), 451–459.

21. Nunez Lopez YO, Garufi G, Seyhan AA. Altered levels of circulating cytokines and microRNAs in lean and obese individuals with pre-diabetes and type 2 diabetes. Mol Biosyst. 2016;13(1), 106–121.

22. Parrizas M, Brugnara L, Esteban Y, et al. Circulating miR-192 and miR-193b are markers of pre-diabetes and are modulated by an exercise intervention. J Clin Endocrinol Metab. 2015;100(3), E407–415.

23. Lee JH, Lee JH, Rane SG. TGF-beta Signaling in Pancreatic Islet beta Cell Development and Function. Endocrinology. 2021;162(3).

24. Hong SH, Kang M, Lee KS, Yu K. High fat diet-induced TGF-beta/Gbb signaling provokes insulin resistance through the tribbles expression. Sci Rep. 2016;6, 30265.

25. Herder C, Zierer A, Koenig W, Roden M, Meisinger C, Thorand B. Transforming growth factor-beta1 and incident type 2 diabetes: results from the MONICA/KORA case-cohort study, 1984-2002. Diabetes Care. 2009;32(10), 1921–1923.

26. Ameling S, Kacprowski T, Chilukoti RK, et al. Associations of circulating plasma microRNAs with age, body mass index and sex in a population-based study. BMC Med Genomics. 2015;8, 61.

